# Use of natural language understanding to facilitate surgical de-escalation of axillary staging in patients with breast cancer

**DOI:** 10.1101/2024.02.03.24302095

**Authors:** Neil Carleton, Gilan Saadawi, Priscilla F. McAuliffe, Atilla Soran, Steffi Oesterreich, Adrian V. Lee, Emilia J. Diego

**Author notes:** **Corresponding Author:** Emilia J. Diego MD FACS, Division of Breast Surgical Oncology, Department of Surgery, University of Pittsburgh School of Medicine, 300 Halket St., Suite 2601, Pittsburgh, PA, 15213.

## Abstract

Natural language understanding (NLU) may be particularly well-equipped for enhanced data capture from the electronic health record (EHR) given its examination of both content- and context-driven extraction. We developed and applied a NLU model to examine rates of pathological node positivity (pN+) and rates of lymphedema to determine if omission of routine axillary staging could be extended to younger patients with ER+/cN0 disease. We found that rates of pN+ and arm lymphedema were similar between patients 55-69yo and ≥70yo, with rates of lymphedema exceeding rates of pN+ for clinical stage T1c and smaller disease. Data from our NLU model suggest that omission of SLNB might be extended beyond Choosing Wisely recommendations, limited to those over 70 years old, to all postmenopausal women with early-stage ER+/cN0 disease. These data support the recently-reported SOUND trial results and provide additional granularity to facilitate surgical de-escalation.

## Introduction

Large language models (LLM) have recently gained attention lately with the inception of widespread use popularized by mainstream media. In general, these LLMs have achieved a paradigm shift in natural language understanding (NLU). Natural language understanding (NLU), rather than processing text as-said, focuses on both content and context of the content, facilitating additional data capture^1^. NLU, like natural language processing (NLP), is particularly well-suited for extraction of key data and details from both structured and unstructured sources from the electronic health record (EHR)^2,3^. At its core, NLU involves both language modeling and reasoning. Language modeling involves the creation of rules and encodings specifically geared toward the features and characteristics of a language architecture. Secondarily, reasoning is another integral task; and although reasoning is not the same as intelligence per se, it involves the necessary process of representing and inferencing a higher value conclusions from a set of information. Use of these cores LLM constructs takes advantage of the machine’s deductive reasoning and ensures validity and consistency. The last decade has seen an exponential increase in the volume of routinely collected data in healthcare. As a result, techniques for handling and interpreting large datasets, including NLU, have become increasingly popular and are now commonly referenced in medical literature^3,4^.

In clinical applications, the advantage of NLU over a human abstracted registry-based approach is the speed and efficiency of data extraction for a large number of patients in real time, as well as the capture of data points not conventionally included in a registry. NLU may be a favorable method for a number of oncologic applications, such as clinical trial matching, identification of lines of systemic treatment, and quality improvement initiatives^5^.

In this report, we evaluate the success of using NLU to determine the rate of pathologically positive lymph nodes (pN+) and rates of lymphedema, comparing these extracted data points between women aged 55-69 years old (middle-aged) against those over 70 (older). Multiples societies, including the NCCN and Society of Surgical Oncology, advocate against the routine use of SLNB for women ≥ 70 years with early-stage ER+/cN0 disease given the low likelihood of axillary involvement and axillary recurrence risk, absence of survival benefit, and greater reliance on genomic testing for therapeutic decisions. While axillary staging impacts adjuvant treatment decision making for many patients, in some cases, the risks may outweigh the harms in terms of breast and arm lymphedema, numbness, and decrements in quality of life following axillary surgery^6,7^. Additionally, in the RxPONDER era, postmenopausal patients with ER+, pN1 disease who have a Recurrence Score ≤ 25 do not benefit from adjuvant chemotherapy, further decreasing utility of axillary staging in this group^8,9^. Therefore, we sought to determine if omission of routine SLNB, a practice adopted for women ≥ 70 with ER+, clinically cN0 breast cancer, may be applicable in younger post-menopausal patients with cN0 disease^10,11^.

## Methods

To this end, all patients with early stage ER+, cN0 breast cancer who had SLNB from January 2015-December 2017 were identified in an integrated academic health network (University of Pittsburgh Medical Center) comprised of 15 hospitals in Western Pennsylvania. This project was approved as a quality improvement initiative by the UPMC Quality Improvement Committee under Project 3618. Patient clinical data were abstracted from the EHR using Realyze Intelligence™ NLU technology. Overall, we processed over 580,000 documents from all inpatient and outpatient records over the time period.

The NLU platform (**Figure 1**) includes a data ingestion and NLP module, which in turn includes an engine and ontological definitions. The engine applies NLP to raw medical data to produce annotated healthcare records, which are conveyed to a reasoning engine. The reasoning engine includes a reasoning engine module which has coupled value sets and data models and rules. These value sets and data models and rules relate to various disease-specific models. The reasoning engine receives the annotated health records as input and analyzes them to generate one or more instances of the disease-specific model(s) for one or more patients. The disease-specific models may include information (e.g., conclusions about the health status of a patient) that were not stated explicitly in the raw medical data. The system may repeat some or all of the process above over time (such as in response to new raw medical data and/or changes in the NLP engine, ontological definitions, value sets, and/or data models/rules) to update the instances of the disease specific models. Previous instances of the disease specific models may be retained by the system.

**Figure 1:**
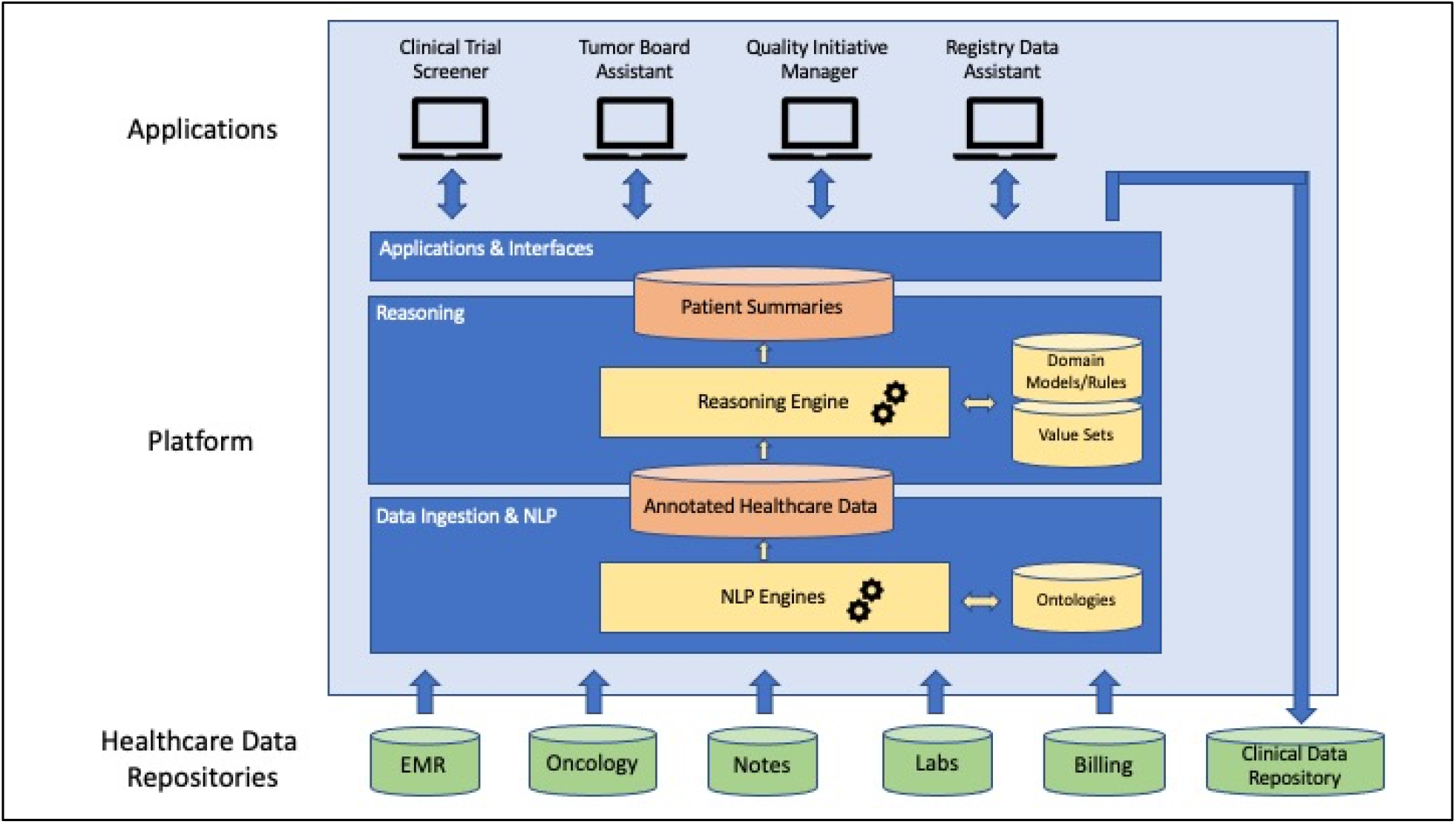
Process of data ingestion and NLP, a reasoning layer, and application to real-world clinical use cases.

**Figure 2:**
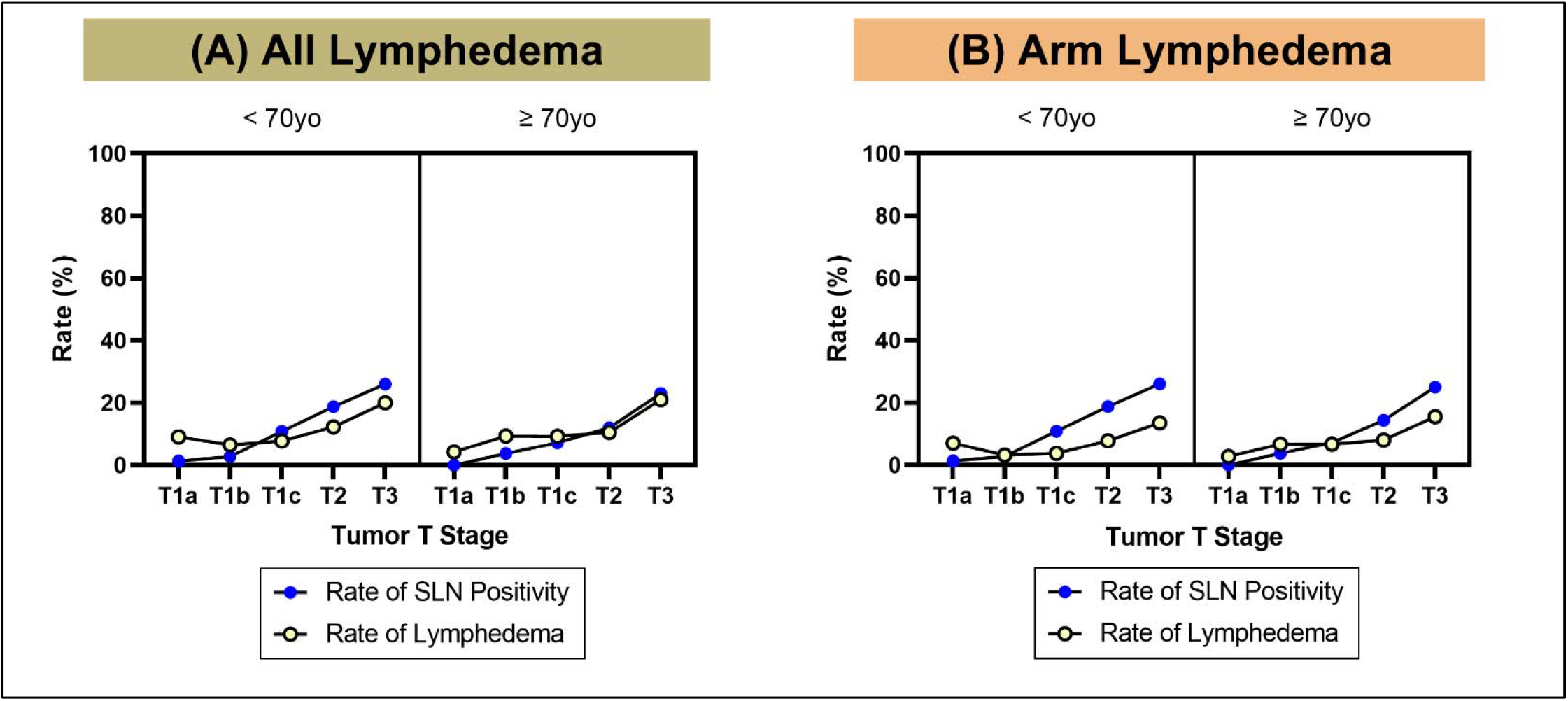
Rates of pathologic node positivity and lymphedema after SLNB for ER+/cN0 disease, broken down by age and stage. The NLU was designed to identify rates of breast and arm lymphedema separately. In (A), we show rates of pN+ along with all lymphedema, which again refers to combined rates of breast and arm lymphedema. In (B), we show pN+ rates overlaid only with arm lymphedema.

The Realyze NLU pipeline uses a combination of LLMs, machine learning algorithms and standard terminologies to create a breast cancer patient model that includes genomic, phenotypic, and clinical data. The pipeline gathers information from all data sources – structured and unstructured – and normalizes the information to create a complete model of patient clinical criteria.

Realyze Information Models use clinical data formatting flexible enough to represent clinical disorders on a concept level as well as the encounter, patient, and population levels. A breast cancer model with focus on the lymph node identification, pathological as well as clinical tumor and node classification, and lymphedema were developed and mapped to standard terminology. A Semantic Reasoning layer is provided by different mechanisms including a rule-based layer to render answers to the questions posed in this hypothesis. The pipeline is modular and the separation between the linguistic NLP from the clinical models allow the rapid refinement of the models without the need of long re-training and allow additions as well as adjustment of the interpretation of data based on clinical guidelines as well as evidence based medicine. In particular for lymphedema identification, we used an iterative approach for rapid model refinement. We found that early versions of the NLU were identifying patients that were being seen for preventative or consultative services, even though they had the I89.0 diagnosis code attached to their medical records. With subsequent iterations, the pre-trained NLU was particularly advantageous in that the model identified context-dependent details that honed in on true, diagnosed lymphedema cases.

NLU performance was validated by manually verifying key clinical variables (i.e., clinical stage, pathologic stage, nodal positivity, and lymphedema) on a subset of patients (random 50% of patients were selected; full review of all clinical details was reviewed for these patients). Statistical analysis to determine any difference in pN+ rates by age was performed using Chi-square testing with significance set at p < 0.05.

## Results

Using the developed NLU approach to extract data from an integrated health system’s EHR, we identified 925 patients age 55 years or older (median age 67.0 years old) diagnosed with early-stage ER+, cN0 breast cancer between 2015 and 2017 period who underwent SLNB. We first evaluated the accuracy of the NLU model. The NLU model yielded strong results for breast and arm lymphedema identification with false positive rates of only 15%, meaning the model identified 85% of patients who correctly had breast and/or arm lymphedema, a key data point for patients with breast cancer and clinicians managing care that is not routinely captured in registry- or population-level databases. False positives, in this instance, largely captured lower extremity lymphedema. Overall, the model accrued 93% per-patient accuracy, where all identified variables, which included stage, grade, Ki-67, ECOG Performance Status, and Oncotype DX Recurrence Scores, and verification of clinical node negativity through a negative ultrasound and clinical exam, were correct.

Comparison of our NLU model against the Cancer Registry, which yielded a comparable number of patients over the designated time period (with median age 67.2 years old) and non-significant differences in rates of pN+ across ages and clinical stages, indicating our model performed well against the registry-based standard. Median follow up time from the date of diagnosis to the date of the last record was 5.5 years.

We then went on to evaluate rates of SLNB positivity and lymphedema. For both middle-aged women and older women, there was an expected increase in pN+ rates as the stage increased (**Figure 3**). When comparing incidence of pN+ disease stratified by age (<70 or ≥70), there was no difference in pN+ rates across all stages (p = 0.80). In particular, equally low rates of SLN positivity were seen for patients across all ages with clinical stage T1a, T1b, and T1c disease, where rates of total lymphedema were higher than rates of pN+. Only one patient in each of age groups had pN2-3 disease, indicating the majority of pN+ cases had 1-3 positive lymph nodes and thus would not receive adjuvant chemotherapy.

## Discussion

This data is in alignment with other published reports on potential omission of SLNB in postmenopausal women with ER+/cN0 disease due to low risk of disease in the axillary nodes^12,13^ and lack of information that would change adjuvant therapy decision making. These data suggest that rates of pN2-3 are uncommon in women with cN0 disease regardless of age. Our data further contribute to this argument by examining rates of lymphedema overlayed with the rates of pN+. Future validation is needed with a larger sample size given the wide variability in de-escalation practices for axillary surgery^14-16^.

The designed NLU model was effective for capturing structured and unstructured data, comparable to the data collected from a registry-based approach. Although pre-training and special model development is necessary, once it is available, NLU may be even more beneficial for extracting information such as lymphedema, where the model was able to correctly distinguish between breast and arm lymphedema and use contextual clues to diminish outputs such as lower extremity lymphedema. Clinicians are uniquely positioned to identify opportunities for NLU to benefit patients, and healthcare systems may employ this new tchnology to better right-size treatments for patients.

Of particular interest is the context of these results in light of the recently-reported SOUND trial, which showed that omission of axillary surgery was noninferior to axillary surgery when evaluating distant disease free survival in patients with cT1N0 breast cancer regardless of age^17^. Our data are concordant with these data, showing that rates of LN positivity remain low in patients with small tumors and suggests axillary surgery in this group does not provide additional information to guide systemic therapy. We provide additional information and granularity in reporting rates of LN positivity and lymphedema, which may further help surgeons and patients in shared decision-making approaches to right-size axillary surgery.

A number of limitations exist when extracting data from the EHR. In this case, our NLU model was limited when it came to extracting data from scanned reports or uploaded images in the medical record, which is common in our health system that attracts patients from non-affiliated hospitals around the region. Secondly, the 5–8-year follow-up of patients may not identify the rare occasion of later-onset lymphedema that can occur decades after SLNB. Lastly, while we did employ an iterative approach for model refinement specifically for lymphedema identification, it stands that there still may be some false positives due to the fact that while lymphedema is the main diagnosis, the patient actually has motion limitations, neuropathy, or other pain syndrome.

In conclusion, these data suggest that the Choosing Wisely recommendation to omit SLNB might be extended to a younger cohort of patients with ER+/cN0 disease, specifically those with stage clinical T1a, T1b, and T1c (those less than 2cm) tumors. With low rates of pN+ disease and less reliance on axillary stage for treatment decision making, the harms of surgical axillary staging, such as risk for lymphedema, may outweigh the benefits. Ongoing clinical trials, such as the BOOG 2013-08 and INSEMA trials, will help to clarify the utility of SLNB in these patient populations.

## Data Availability

All data produced in the present study are available upon reasonable request to the authors.

## ACKNOWLEDGEMENTS

We acknowledge the many contributions of the patients, families, researchers, clinical staff, and sponsors of this study. We thank Sharon Winters & Vonda Mazzarella from the UPMC Cancer Registry for their help with this report.

## Author Contributions

Concept and Design: AVL, EJD, GS

Acquisition, Analysis, or Interpretation of the Data: EJD, NC, GS, AS

Drafting the Manuscript: NC, AVL, EJD, SO, PFM

Critical Revision of the Manuscript: NC, AVL, PFM, EJD, GS, SO, AS

Statistical Analysis: NC, GS, AVL, EJD

Obtained Funding: AVL, SO, NC

Administrative, Technical, or Material Support: AVL, SO, PFM, EJD

Supervision: AVL, SO, PFM, EJD

## Data Availability Statement

The data underlying this article cannot be shared due to the fact that it contains identifiable patient information. Additional summary level data without individual data may be available upon request to the corresponding author (EJD).

